# Participant Outcomes of the Diabetes and WELLbeing 12-Week Psychoeducational Self-Management Programme for Type 2 Diabetes: A Four-Country European Evaluation

**DOI:** 10.64898/2026.11.13.26368046

**Authors:** Anurup Mukherjee, Sukhi S. Shergill, Sarah Hotham, Ruben Vanbosseghem, Maarten Gijssel, Sharon Manship, Sabina Hulbert, Eleni Hatzidimitriadou

## Abstract

**Introduction:** Diabetes self-management education and support (DSMES) improves glycaemic control, but its benefits often fade once programmes end, and psychosocial outcomes are evaluated less often. We evaluated participant outcomes of the Diabetes and WELLbeing (DWELL) programme, a 12-week psychoeducational self-management programme for adults with type 2 diabetes delivered in four European countries.

**Methods:** In this uncontrolled pre-post evaluation, 605 adults with type 2 diabetes were recruited in the UK, France, Belgium and the Netherlands. Outcomes were assessed at baseline, at programme completion (12 weeks) and at 6-month follow-up. The primary outcome was diabetes self-care (Summary of Diabetes Self-Care Activities); secondary outcomes included HbA1c, weight, BMI, waist circumference, empowerment, illness perceptions, eating behaviour, health status, physical activity and quality of life. Within-person changes were tested with paired Wilcoxon signed-rank tests.

**Results:** Of 605 participants, 453 were assessed at programme completion and 293 at 6 months. Most self-care behaviours improved by programme completion (for example, dietary self-care rose from a median of 2 to 3 days per week), and all seven self-care measures differed significantly from baseline at 6 months (r=0.41 to 0.82; all p<0.001). By programme completion, weight (median 1.25 kg), waist circumference (2.0 cm) and HbA1c (2.5 mmol/mol) had fallen, and empowerment, illness coherence, emotional and external eating, physical and mental health and quality of life had improved (all p<0.001). At 6 months, all metabolic outcomes remained below baseline, although HbA1c had partly rebounded (1.7 mmol/mol below baseline), and most psychosocial gains were sustained.

**Conclusions:** Participation in DWELL was associated with improvements in self-care, metabolic and psychosocial outcomes that were partly sustained 6 months after the programme ended. As an uncontrolled evaluation, the study cannot establish causation, and controlled studies with longer follow-up are needed.

**Trial registration:** ClinicalTrials.gov NCT04274660 (registered retrospectively in February 2020).

## Introduction

Diabetes is among the ten leading causes of death and disability worldwide, responsible for an estimated 1.5 million deaths each year and a major contributor to cardiovascular disease, kidney failure, visual impairment and lower-limb amputation.^1^ Type 2 diabetes mellitus (T2DM) accounts for around 90% of all diabetes cases, typically presenting later in life, and its prevalence has risen sharply across countries of every income level over the past three decades, including across Europe.^1–3^ Global diabetes-related health expenditure reached an estimated US$966 billion in 2021,^3^ a cost that falls heavily on individuals, families and health systems alike.^4^ This growth reflects a complex mix of socioeconomic, demographic, environmental and genetic factors, including an ageing population, urbanisation, physical inactivity and rising rates of overweight and obesity.^3^

T2DM is a lifelong condition, and much of the day-to-day work of managing it falls to people with diabetes and their families, which is why effective self-management sits at the heart of good diabetes care. Diabetes self-management education and support (DSMES) aims to give people the knowledge, practical skills and confidence to manage their condition in partnership with healthcare professionals,^5^ and it reduces glycated haemoglobin (HbA1c) relative to usual care, with combined education and support outperforming education alone.^6,7^ These benefits, however, tend to be greatest during active programme contact and to fade once it ends,^8,9^ and DSMES has accordingly evolved from a time-limited, knowledge-focused intervention towards a more continuous model that pairs education with behavioural, emotional and practical support.^5,6^

Despite wide agreement on its value, there is no single agreed model of DSMES delivery: programmes differ in duration, content, setting and the extent of individual tailoring, and provision varies considerably across European health systems.^10^ Diabetes education has also moved away from a compliance-based, knowledge-oriented model towards one built around empowerment, shared decision-making and self-management,^11^ and self-efficacy, perceived control and illness understanding all appear to shape whether knowledge translates into everyday self-management. Yet psychological outcomes are assessed far less often than metabolic ones in evaluations of DSME,^11^ so broader evaluation is needed that examines empowerment, illness perceptions, emotional wellbeing, eating behaviour and self-care alongside the usual biomedical measures.

The Diabetes and WELLbeing (DWELL) project was developed with these interconnected needs in mind. DWELL was a 12-week psychoeducational self-management programme for adults with T2DM, co-designed by people living with diabetes, family carers, healthcare professionals and academics. It brought together four core strands: diabetes education, nutrition, physical activity and wellbeing. It drew on motivational interviewing, peer support and flexible activities shaped around participants’ own goals and circumstances. The programme was developed and evaluated in four European countries following the Medical Research Council framework for complex interventions.^12^ This paper reports the quantitative participant outcomes from that evaluation, structured around the study’s registered outcomes and addressing two research questions:

1. What was the impact of the DWELL programme on diabetes self-care, the study’s primary outcome?
2. What was its impact on the secondary outcomes of health status, illness perception, empowerment, eating behaviour, and the metabolic measures (BMI, waist circumference and HbA1c)?

## Materials and Methods

### Study design and ethical approval

The evaluation used a quasi-experimental, non-randomised, pre–post design nested within a longitudinal mixed-methods study. Outcomes were measured at baseline (T0), at the end of the 12-week programme (T1), and at 6-month (T2) and 12-month (T3) follow-up. Quantitative data were collected using the DWELL Tool, a set of validated questionnaires chosen to address the study’s research questions, alongside a form recording demographic, biomedical and metabolic information. Qualitative data came from end-of-programme focus groups, semi-structured interviews and researcher field notes. This paper reports the quantitative findings from baseline (T0), programme completion (T1) and the first follow-up point (T2). Data collection at the 12-month follow-up (T3) was substantially disrupted by the COVID-19 pandemic and associated public health measures, which affected sites in all four countries. Shielding advice for people with T2DM, the suspension of group-based delivery and the interruption of face-to-face assessment affected cohorts unevenly, leaving T3 data too incomplete and too unevenly distributed between sites and cohorts to support meaningful analysis. T3 outcomes are therefore not reported here, and the 6-month follow-up represents the longest interval for which comparable data are available across the full sample.

Approval was obtained at each participating site. In the UK, the London Bridge Research Ethics Committee approved the study (REC reference 17/LO/1480), with Canterbury Christ Church University as sponsor. In France, the Comité de Protection des Personnes Nord Ouest IV gave a favourable opinion on 29 March 2018 (CPP reference 17/36; national registration number 2017-A00720-53), with Centre Hospitalier de Douai as sponsor. In Belgium, the Medical Ethics Committee of Ghent University Hospital approved the study on 22 August 2018 (reference B670201836754). In the Netherlands, the Medisch Ethische Toetsingscommissie Brabant confirmed on 20 July 2017 that the study was not subject to the Medical Research Involving Human Subjects Act, and the requirement for formal approval was therefore waived (reference NL62544.028.17). All participants gave written informed consent before taking part. The study was conducted in accordance with the Declaration of Helsinki.

### Participants

Adults aged 18 or over with a diagnosis of T2DM were eligible. In total, 605 participants were recruited across the UK (n=299), France (n=203), the Netherlands (n=55) and Belgium (n=48). Participants ranged in age from 25 to 93 years (mean 62.5, SD 10.0). Among those with recorded sex data, 280 (47.2%) identified as male and 313 (52.8%) as female. Most participants (539, 92.0%) reported their ethnicity as White; 21 (3.5%) as Asian; 15 (2.5%) as Black, African or Caribbean; 8 (1.3%) as mixed ethnicity; and 3 (0.5%) as another ethnicity. Baseline characteristics are summarised in Table 1.

**Table 1.**
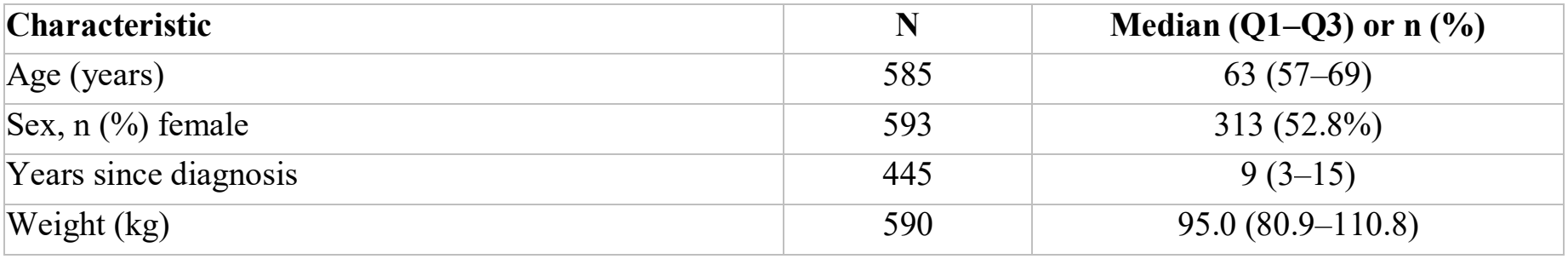
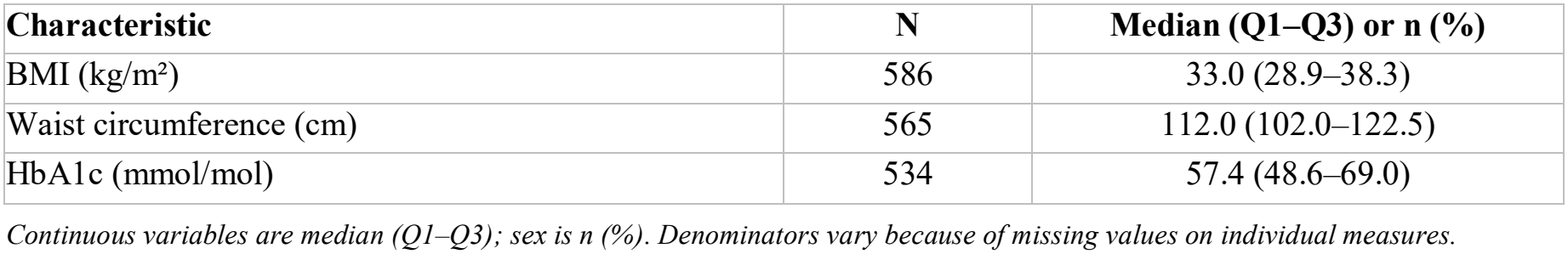
Baseline (T0) sample characteristics.

| **Characteristic** | **N** | **Median (Q1–Q3) or n (%)** |
|---|---|---|
| Age (years) | 585 | 63 (57–69) |
| Sex, n (%) female | 593 | 313 (52.8%) |
| Years since diagnosis | 445 | 9 (3–15) |
| Weight (kg) | 590 | 95.0 (80.9–110.8) |
| **Characteristic** | **N** | **Median (Q1–Q3) or n (%)** |
| BMI (kg/m²) | 586 | 33.0 (28.9–38.3) |
| Waist circumference (cm) | 565 | 112.0 (102.0–122.5) |
| HbA1c (mmol/mol) | 534 | 57.4 (48.6–69.0) |

*Continuous variables are median (Q1–Q3); sex is n (%). Denominators vary because of missing values on individual measures.*

**Table 2.**
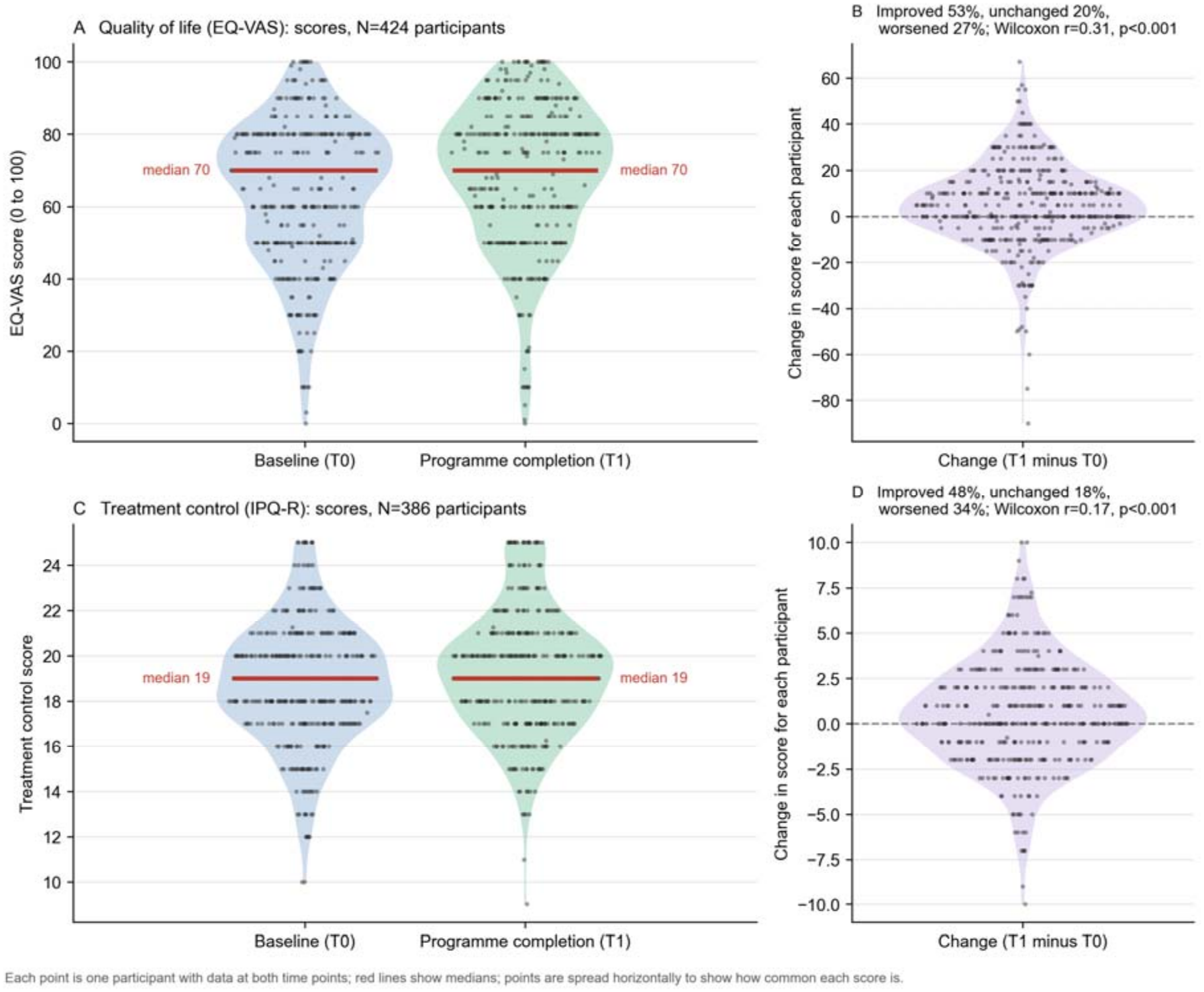
Participant-reported outcomes and assessment instruments.

| Panel | Text |
|---|---|
| A | Quality of life (EQ-VAS): scores, N=424 participants |
| B | Improved 53%, unchanged 20%,<br>worsened 27%; Wilcoxon r=0.31, p<0.001 |
| C | Treatment control (IPQ-R): scores, N=386 participants |
| D | Improved 48%, unchanged 18%,<br>worsened 34%; Wilcoxon r=0.17, p<0.001 |

Each point is one participant with data at both time points; red lines show medians; points are spread horizontally to show how common each score is.

Numbers contributing data at each subsequent time point were lower, and varied between outcomes, because of attrition, missing or incomplete responses to individual measures, and outcome-specific eligibility (for example, insulin adherence was assessed only among participants prescribed insulin). Medians at each time point were therefore based on all available data, whereas each within-person comparison included only participants with data at both of the time points being compared; the number contributing to each comparison is reported alongside every result in Tables 3 and 4 and in the accompanying text. Retention varied by country (see Participant flow), but participants retained at follow-up were comparable to those lost on most baseline characteristics (see Comparison of participants retained at follow-up with those lost).

**Table 3.**
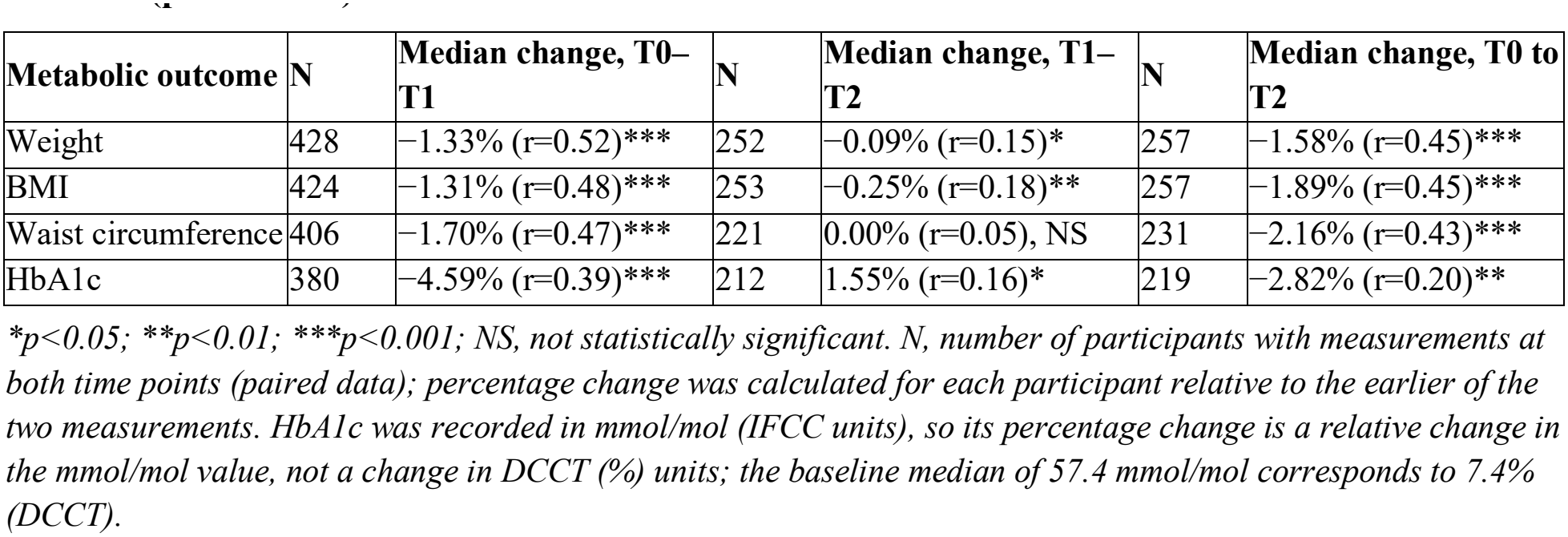
Median percentage changes in metabolic outcomes from T0 to T1, from T1 to T2 and from T0 to T2 (paired data)

| **Metabolic outcome** | **N** | **Median change, T0–T1** | **N** | **Median change, T1–T2** | **N** | **Median change, T0 to T2** |
|---|---|---|---|---|---|---|
| Weight | 428 | −1.33% (r=0.52)*** | 252 | −0.09% (r=0.15)* | 257 | −1.58% (r=0.45)*** |
| BMI | 424 | −1.31% (r=0.48)*** | 253 | −0.25% (r=0.18)** | 257 | −1.89% (r=0.45)*** |
| Waist circumference | 406 | −1.70% (r=0.47)*** | 221 | 0.00% (r=0.05), NS | 231 | −2.16% (r=0.43)*** |
| HbA1c | 380 | −4.59% (r=0.39)*** | 212 | 1.55% (r=0.16)* | 219 | −2.82% (r=0.20)** |

*\*p<0.05; \*\*p<0.01; \*\*\*p<0.001; NS, not statistically significant. N, number of participants with measurements at both time points (paired data); percentage change was calculated for each participant relative to the earlier of the two measurements. HbA1c was recorded in mmol/mol (IFCC units), so its percentage change is a relative change in the mmol/mol value, not a change in DCCT (%) units; the baseline median of 57.4 mmol/mol corresponds to 7.4% (DCCT).*

**Table 4.**
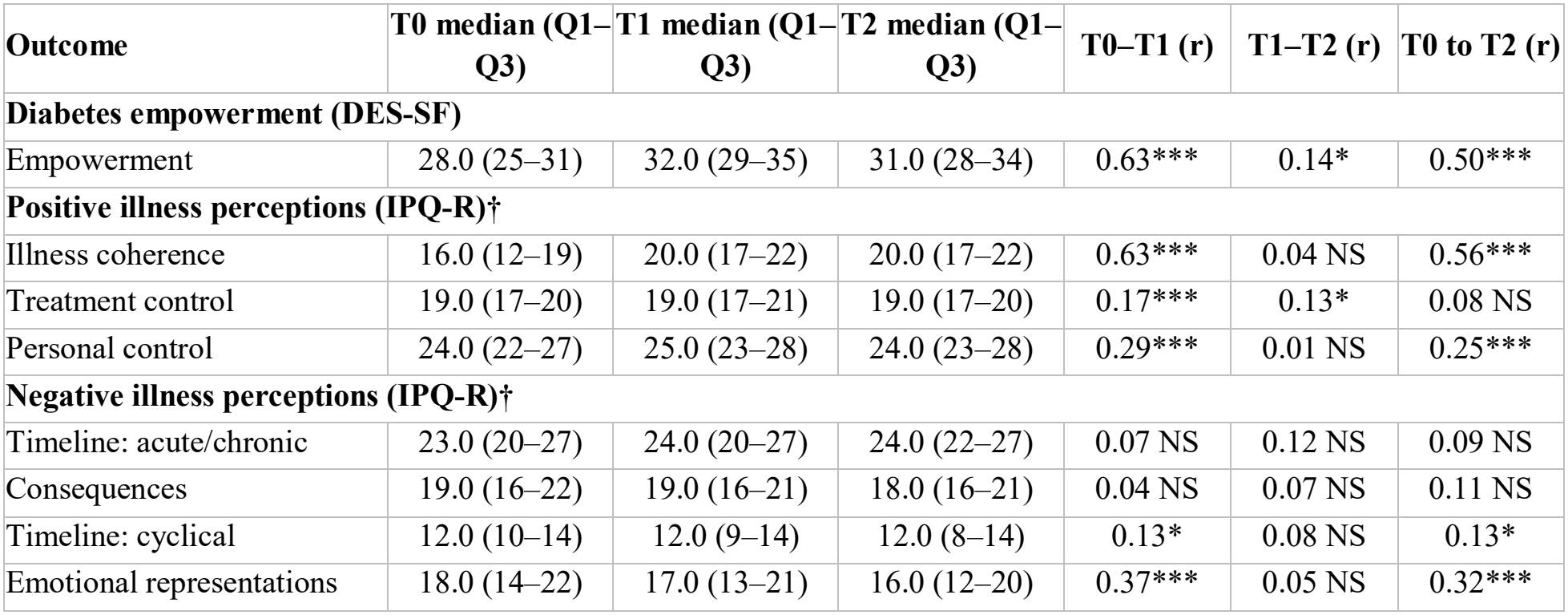
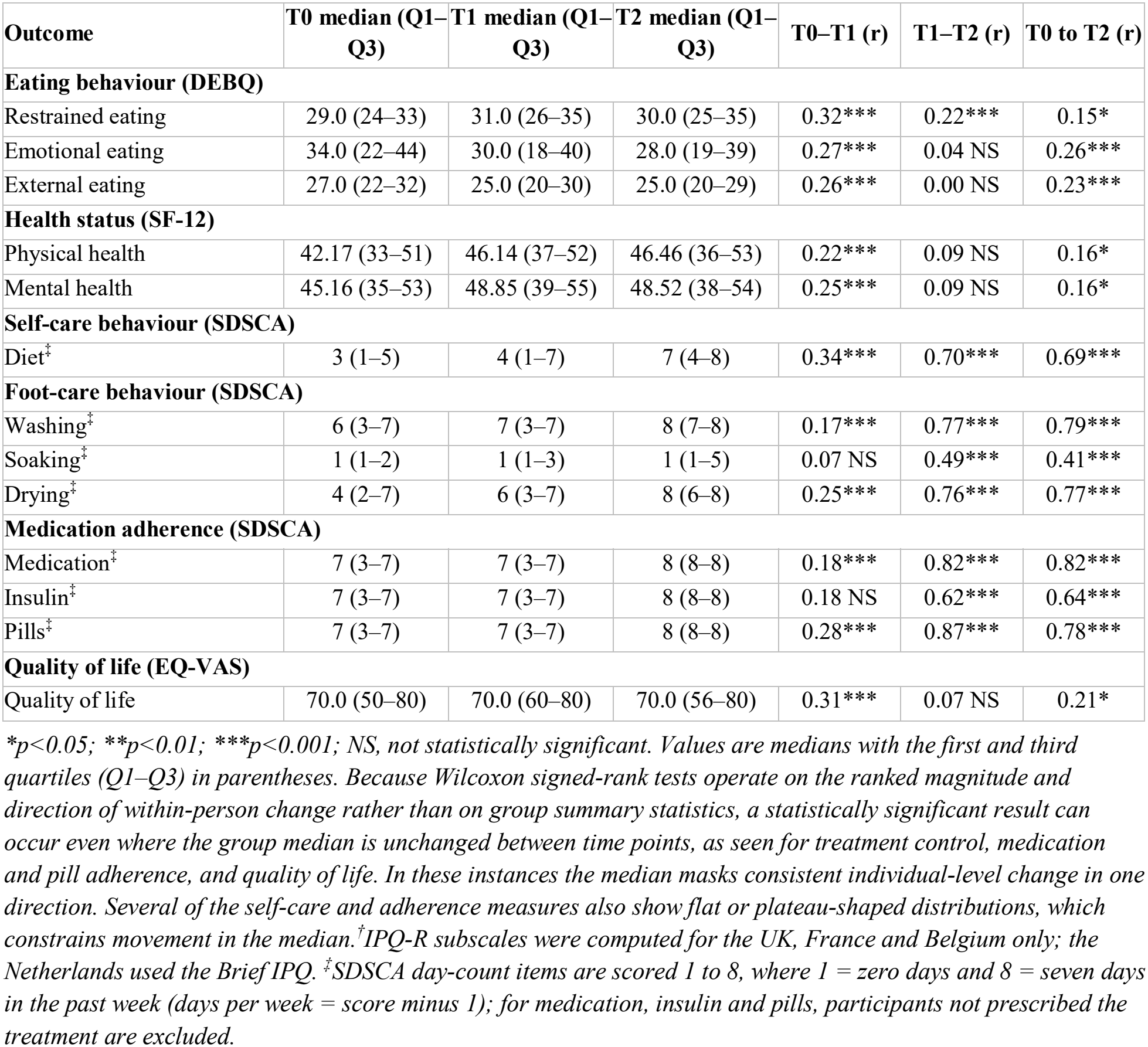
Median (Q1–Q3) psychological and self-management outcome scores at T0, T1 and T2, with effect sizes.

| **Outcome** | **T0 median (Q1–Q3)** | **T1 median (Q1–Q3)** | **T2 median (Q1–Q3)** | **T0–T1 (r)** | **T1–T2 (r)** | **T0 to T2 (r)** |
|---|---|---|---|---|---|---|
| **Diabetes empowerment (DES-SF)** |  |  |  |  |  |  |
| Empowerment | 28.0 (25–31) | 32.0 (29–35) | 31.0 (28–34) | 0.63*** | 0.14* | 0.50*** |
| **Positive illness perceptions (IPQ-R)†** |  |  |  |  |  |  |
| Illness coherence | 16.0 (12–19) | 20.0 (17–22) | 20.0 (17–22) | 0.63*** | 0.04 NS | 0.56*** |
| Treatment control | 19.0 (17–20) | 19.0 (17–21) | 19.0 (17–20) | 0.17*** | 0.13* | 0.08 NS |
| Personal control | 24.0 (22–27) | 25.0 (23–28) | 24.0 (23–28) | 0.29*** | 0.01 NS | 0.25*** |
| **Negative illness perceptions (IPQ-R)†** |  |  |  |  |  |  |
| Timeline: acute/chronic | 23.0 (20–27) | 24.0 (20–27) | 24.0 (22–27) | 0.07 NS | 0.12 NS | 0.09 NS |
| Consequences | 19.0 (16–22) | 19.0 (16–21) | 18.0 (16–21) | 0.04 NS | 0.07 NS | 0.11 NS |
| Timeline: cyclical | 12.0 (10–14) | 12.0 (9–14) | 12.0 (8–14) | 0.13* | 0.08 NS | 0.13* |
| Emotional representations | 18.0 (14–22) | 17.0 (13–21) | 16.0 (12–20) | 0.37*** | 0.05 NS | 0.32*** |
| **Outcome** | **T0 median (Q1–Q3)** | **T1 median (Q1–Q3)** | **T2 median (Q1–Q3)** | **T0–T1 (r)** | **T1–T2 (r)** | **T0 to T2 (r)** |
| **Eating behaviour (DEBQ)** |  |  |  |  |  |  |
| Restrained eating | 29.0 (24–33) | 31.0 (26–35) | 30.0 (25–35) | 0.32*** | 0.22*** | 0.15* |
| Emotional eating | 34.0 (22–44) | 30.0 (18–40) | 28.0 (19–39) | 0.27*** | 0.04 NS | 0.26*** |
| External eating | 27.0 (22–32) | 25.0 (20–30) | 25.0 (20–29) | 0.26*** | 0.00 NS | 0.23*** |
| **Health status (SF-12)** |  |  |  |  |  |  |
| Physical health | 42.17 (33–51) | 46.14 (37–52) | 46.46 (36–53) | 0.22*** | 0.09 NS | 0.16* |
| Mental health | 45.16 (35–53) | 48.85 (39–55) | 48.52 (38–54) | 0.25*** | 0.09 NS | 0.16* |
| **Self-care behaviour (SDSCA)** |  |  |  |  |  |  |
| Diet<sup>‡</sup> | 3 (1–5) | 4 (1–7) | 7 (4–8) | 0.34*** | 0.70*** | 0.69*** |
| **Foot-care behaviour (SDSCA)** |  |  |  |  |  |  |
| Washing<sup>‡</sup> | 6 (3–7) | 7 (3–7) | 8 (7–8) | 0.17*** | 0.77*** | 0.79*** |
| Soaking<sup>‡</sup> | 1 (1–2) | 1 (1–3) | 1 (1–5) | 0.07 NS | 0.49*** | 0.41*** |
| Drying<sup>‡</sup> | 4 (2–7) | 6 (3–7) | 8 (6–8) | 0.25*** | 0.76*** | 0.77*** |
| **Medication adherence (SDSCA)** |  |  |  |  |  |  |
| Medication<sup>‡</sup> | 7 (3–7) | 7 (3–7) | 8 (8–8) | 0.18*** | 0.82*** | 0.82*** |
| Insulin<sup>‡</sup> | 7 (3–7) | 7 (3–7) | 8 (8–8) | 0.18 NS | 0.62*** | 0.64*** |
| Pills<sup>‡</sup> | 7 (3–7) | 7 (3–7) | 8 (8–8) | 0.28*** | 0.87*** | 0.78*** |
| **Quality of life (EQ-VAS)** |  |  |  |  |  |  |
| Quality of life | 70.0 (50–80) | 70.0 (60–80) | 70.0 (56–80) | 0.31*** | 0.07 NS | 0.21* |

*\*p<0.05; \*\*p<0.01; \*\*\*p<0.001; NS, not statistically significant. Values are medians with the first and third quartiles (Q1–Q3) in parentheses. Because Wilcoxon signed-rank tests operate on the ranked magnitude and direction of within-person change rather than on group summary statistics, a statistically significant result can occur even where the group median is unchanged between time points, as seen for treatment control, medication and pill adherence, and quality of life. In these instances the median masks consistent individual-level change in one direction. Several of the self-care and adherence measures also show flat or plateau-shaped distributions, which constrains movement in the median.<sup>†</sup>IPQ-R subscales were computed for the UK, France and Belgium only; the Netherlands used the Brief IPQ. <sup>‡</sup>SDSCA day-count items are scored 1 to 8, where 1 = zero days and 8 = seven days in the past week (days per week = score minus 1); for medication, insulin and pills, participants not prescribed the treatment are excluded.*

### Intervention

Participants could self-refer or be referred by GPs, hospitals or other healthcare professionals involved in their care. The development and practical delivery of DWELL have been described in detail elsewhere.^13^

In brief, DWELL was a 12-week psychoeducational programme built around four core areas of diabetes education, nutrition, physical activity and wellbeing. Alongside the core sessions, participants were signposted to optional “pick and mix” activities outside the formal programme, such as local gym sessions or yoga classes; some were free to participants (for example, walking groups set up in France), while others were at participants’ own cost. The programme drew on self-management theory, peer support, motivational interviewing and principles of adult learning, allowing content and activities to be adapted to participants’ circumstances, priorities and readiness for change.

### Methods of assessment

#### Patient-reported outcomes

At each assessment point, participants completed validated measures of diabetes self-care (the primary outcome), physical and mental health, physical activity, illness perceptions, empowerment, eating behaviour and health-related quality of life. These instruments are summarised in Table 2.

#### Metabolic outcomes

Metabolic outcomes comprised body weight (kg), waist circumference (cm), BMI and HbA1c. HbA1c was measured from finger-prick capillary blood samples by trained staff, using the Quo-Test HbA1c Analyzer; in Belgium it was obtained by venous blood draw at the participant’s GP. Changes are expressed as percentage change scores, to account for variation in baseline values and to allow comparison across outcomes measured on different scales.^21,22^

### Statistical analysis

The study was registered with ClinicalTrials.gov (NCT04274660); registration was completed in February 2020, after recruitment had begun in September 2017, but before data collection was complete and before any of the analyses reported here were undertaken. The registered primary outcome was diabetes self-care (SDSCA); the secondary outcomes were health status (SF-12), physical activity (IPAQ), illness perception (IPQ-R), diabetes empowerment (DES-SF), eating behaviour (DEBQ), BMI, waist circumference and HbA1c. Health-related quality of life was assessed using the EQ-VAS from the EQ-5D-3L; the EQ-5D-3L utility index and quality-adjusted life years are not reported here, as the non-intervention comparison group was not recruited owing to the COVID-19 pandemic. This paper reports the within-participant (pre-post) outcomes of the DWELL participants, using paired comparisons of each participant’s measurements between time points.

Distributional assumptions were assessed before analysis. The metabolic variables were strongly right-skewed and heavy-tailed (weight, BMI and HbA1c: skewness +1.2 to +1.4; excess kurtosis +2.0 to +7.4). Several self-care and adherence measures showed moderate left skew or flat, plateau-shaped distributions, and quality of life and medication adherence clustered towards the upper end of their ranges. Formal testing indicated departure from normality for nearly all outcomes (Shapiro–Wilk p<0.05), though with a sample of this size such tests are highly sensitive to minor deviations. Given the pronounced skew in the metabolic outcomes, non-parametric methods were used for the primary unadjusted within-person comparisons; supplementary covariate-adjusted analyses used linear regression with participant-clustered standard errors. Percentage changes in metabolic outcomes between T0 and T1, between T1 and T2, and between T0 and T2, were tested using one-sample Wilcoxon signed-rank tests, with effect sizes reported as r (calculated as Z/√N; values of 0.1, 0.3 and 0.5 were interpreted as small, moderate and large effects, respectively). Linear regression was used to examine whether demographic characteristics predicted percentage change in weight, HbA1c and waist circumference from T0 to T1.

Changes in patient-reported outcomes between T0 and T1, between T1 and T2, and between T0 and T2, were similarly tested using paired Wilcoxon signed-rank tests. For the multidimensional instruments, subscale scores rather than total scores were analysed: the IPQ-R was analysed by individual domain (illness coherence, treatment control, personal control, timeline acute/chronic, consequences, timeline cyclical and emotional representations), the DEBQ by its restrained, emotional and external eating subscales, the SF-12 by its physical and mental component summary scores, and the SDSCA by its dietary, foot-care and medication-adherence items. The DES-SF and EQ-VAS were analysed as single summary scores. SDSCA day-count items were scored on the DWELL coding framework’s 1–8 scale (1 = zero days, 8 = seven days), applied consistently across all time points. For the medication, insulin and pill-taking items, responses of “not applicable” (no such medication prescribed) were treated as missing, so that adherence was assessed only among participants prescribed the relevant medication; denominators are therefore smaller for these items, most notably insulin. Statistical significance was set at p<0.05.

Medians at each time point were based on all available data, and each paired comparison included participants with data at both of the time points being compared.

No correction for multiple comparisons was applied to the longitudinal outcome comparisons; Benjamini–Hochberg correction was applied separately to the retained-versus-lost baseline comparisons (see below). Given the number of outcomes and the repeated comparisons across time points, the statistically significant longitudinal findings should be interpreted with appropriate caution and regarded as hypothesis-generating rather than confirmatory. This is particularly relevant for the smaller effects, where statistical significance largely reflects the size of the sample rather than the magnitude of change. The primary analyses were unadjusted; to test whether the programme effects were robust to participant characteristics, a supplementary set of covariate-adjusted models was also fitted (Supplementary Table S1).

## Results

Of the 605 participants assessed at baseline, the number contributing data at each subsequent time point varied by outcome. For the metabolic outcomes, between 380 and 428 participants contributed data to the T0–T1 comparison and between 212 and 253 to the T1–T2 comparison, and between 219 and 257 to the T0 to T2 comparison (Table 3). For the patient-reported outcomes, between 297 and 428 participants contributed to each T0 to T1 comparison, between 117 and 264 to each T1 to T2 comparison and between 137 and 267 to each T0 to T2 comparison; numbers were smallest for insulin adherence (89, 39 and 45, respectively), which was assessed only among participants prescribed insulin. The denominator for each comparison is reported alongside the corresponding result throughout.

### Participant flow

Figure 1 shows the flow of participants from enrolment to the six-month follow-up. Of 605 participants enrolled, 453 were assessed at the end of the programme and 293 at six-month follow-up. Retention differed markedly between countries: it was lowest in Belgium (8 of 48 retained at follow-up), where COVID-19 interrupted delivery at all three sites; delivery did not resume after the pandemic. This pattern accounts for a substantial share of the overall attrition. Because participants were affected at different stages of their programme and to differing degrees between countries, the pandemic represents an unmeasured source of heterogeneity. Counts are based on the presence of any outcome data at each time point.

**Figure 1.**
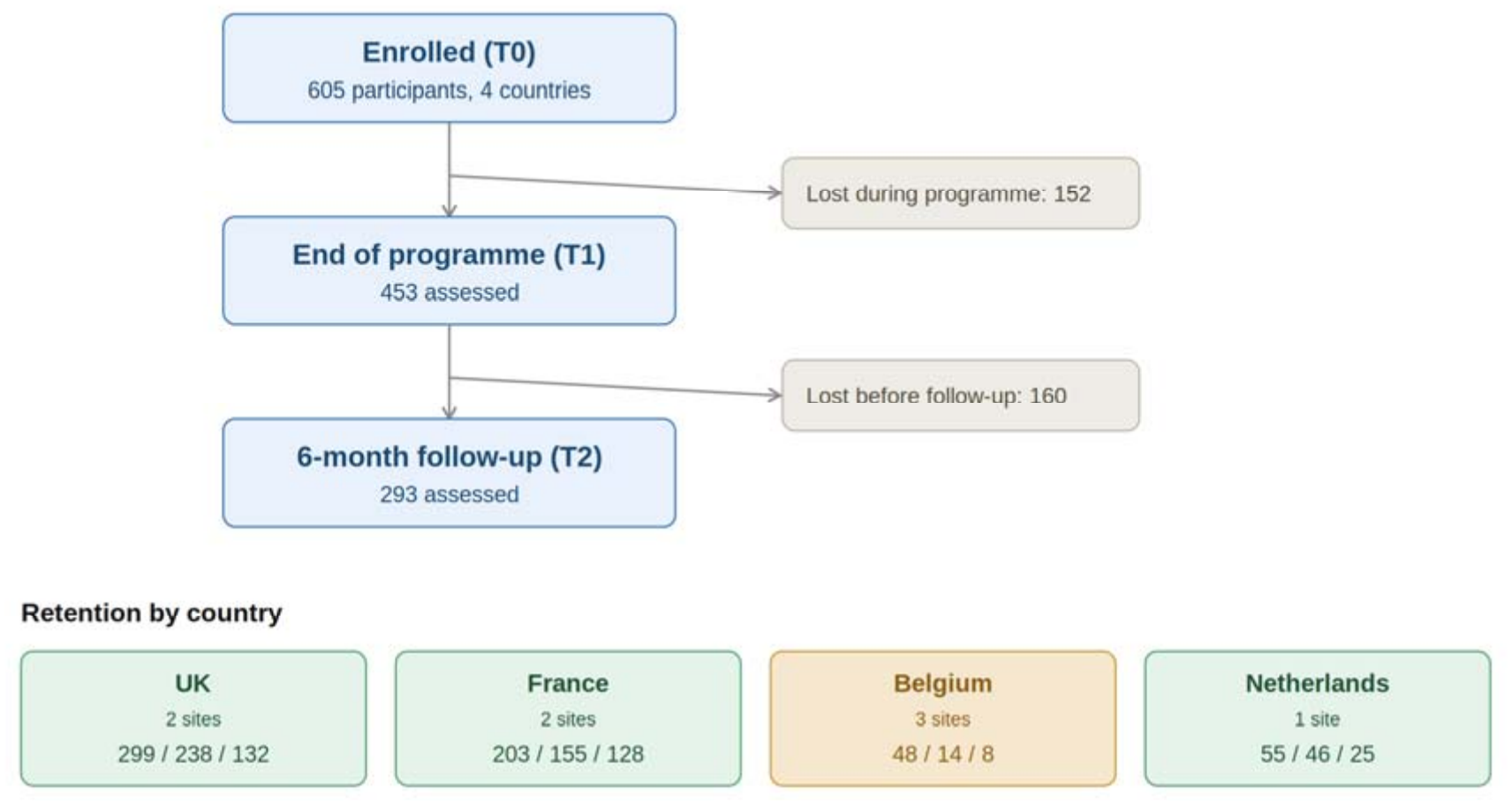
Participant-flow diagram from enrolment to six-month follow-up. *Each country shows T0 / T1 / T2 participant counts, with the number of delivery sites shown beneath the country name. Belgium (highlighted) shows the steepest attrition, following COVID-19 disruption to delivery*.

### Primary outcome: diabetes self-care (SDSCA)

Dietary self-care, measured as the number of days in the past week on which carbohydrate intake was spaced evenly through the day, increased from a median of 2 days at baseline to 3 days at programme completion (N=398, r=0.34, p<0.001) and to 6 days at six-month follow-up (N=153, r=0.70, p<0.001).

Foot care also improved. The median number of days per week on which participants washed their feet rose from 5 at baseline to 6 at programme completion (N=428, r=0.17, p<0.001) and to 7 at follow-up (N=154, r=0.77, p<0.001), and drying between the toes rose from 3 to 5 days (N=426, r=0.25, p<0.001) and then to 7 days (N=154, r=0.76, p<0.001). Foot soaking, which foot-care advice generally discourages, remained uncommon (median 0 days at each time point) and did not change significantly by programme completion (N=414, r=0.07), although it became more frequent by follow-up (N=150, r=0.49, p<0.001).

Among participants prescribed each treatment, median adherence was 6 of 7 days at baseline and at programme completion, equivalent to taking medication as recommended on about 86% of days, rising to 7 of 7 days (100%) at follow-up. Although the medians were unchanged between baseline and programme completion, adherence increased significantly over that period for prescribed medication overall (N=368, r=0.18, p<0.001) and for tablets (N=297, r=0.28, p<0.001), reflecting small but consistent individual-level gains, whereas insulin adherence did not change significantly (N=89, r=0.18). All three measures increased further by follow-up (medication N=137, r=0.82; tablets N=117, r=0.87; insulin N=39, r=0.62; all p<0.001). Because the SDSCA records the number of days on which medication was taken as recommended rather than the proportion of prescribed doses, these figures approximate, but are not equivalent to, dose-based measures of adherence.

Compared with baseline, all seven self-care measures differed significantly at six-month follow-up (r=0.41 to 0.82, all p<0.001; Table 4).

#### Secondary outcomes

#### Effect of the DWELL programme on metabolic health

Table 3 shows the median percentage change in weight, BMI, waist circumference and HbA1c from baseline to programme completion (T0–T1), from programme completion to six-month follow-up (T1– T2), and from baseline to six-month follow-up (T0 to T2), with corresponding effect sizes.

By programme completion, all four metabolic outcomes had improved significantly. The largest effect sizes were seen for weight and BMI, followed by waist circumference and then HbA1c. In original units, these were median reductions of 1.25 kg in weight, 0.4 kg/m^2^ in BMI, 2.0 cm in waist circumference and 2.5 mmol/mol in HbA1c.

Between programme completion and six-month follow-up, weight and BMI continued to fall slightly; these changes were statistically significant but small in magnitude, with correspondingly small effect sizes, and their clinical significance is therefore uncertain. Waist circumference showed no significant change. HbA1c, by contrast, rose by a median of 1.55% (relative to its value at programme completion) over this period, indicating a partial reversal of the improvement achieved during the programme.

Compared with baseline, all four metabolic outcomes remained significantly lower at six-month follow-up (T0 to T2; Table 3): weight by a median of 1.58% (1.4 kg; r=0.45), BMI by 1.89% (0.6 kg/m^2^; r=0.45) and waist circumference by 2.16% (2.0 cm; r=0.43), all p<0.001, and HbA1c by 2.82% (1.7 mmol/mol; r=0.20, p=0.004). HbA1c therefore remained below its baseline level despite rising after the programme ended.

BMI was not modelled separately because of its mathematical dependence on weight. Exploratory models therefore examined percentage change in weight, HbA1c and waist circumference as three distinct outcomes, with HbA1c the more discerning indicator of glycaemic control and waist circumference a marker of metabolic risk independent of glucose. These outcomes were also carried into the covariate-adjusted models.

#### Demographic predictors of change

Participant characteristics recorded at baseline comprised age, sex, ethnicity, country of participation and time since diagnosis. Each was examined as a candidate predictor of percentage change in weight, HbA1c and waist circumference from baseline to programme completion. No characteristic was meaningfully predictive: time since diagnosis, the strongest candidate, explained no more than 1.4% of the variance in change for any of the three measures (weight R^2^=0.013, p=0.040; HbA1c R^2^=0.006, p=0.190; waist R^2^=0.014, p=0.034). Age, sex, ethnicity and country were not significant predictors of weight change. Because baseline characteristics did not predict individual response, these exploratory models are not reported in detail; the covariate-adjusted analyses (Supplementary Table S1) provide a more appropriate treatment of demographic and country effects; in these models, the programme effects on weight, waist circumference, HbA1c, empowerment and physical health all persisted after adjustment for age, sex, time since diagnosis, country and baseline physical health.

#### Effect of the DWELL programme on psychological and self-management outcomes

Table 4 summarises changes across the psychological and self-management measures from T0 to T1, from T1 to T2 and from T0 to T2.

Several outcomes in Table 4 show the same median at successive time points despite a statistically significant change. This is because the Wilcoxon signed-rank test assesses whether individual participants’ scores tend to move in the same direction, not whether the group median shifts. On scales with coarse response steps, many participants can improve by a small amount while the score of the middle-ranked participant stays the same, so the median does not move. Quality of life illustrates this most clearly, with a median of 70 at all three time points but a significant improvement from T0 to T1 (r=0.31); treatment control shows the same pattern, its median unchanged at 19 despite a significant change. Figure 2 illustrates this for both outcomes: neither median moved, but more participants improved than worsened (quality of life 53% versus 27%, with a median individual change of +5 points; treatment control 48% versus 34%).

**Figure 2.**
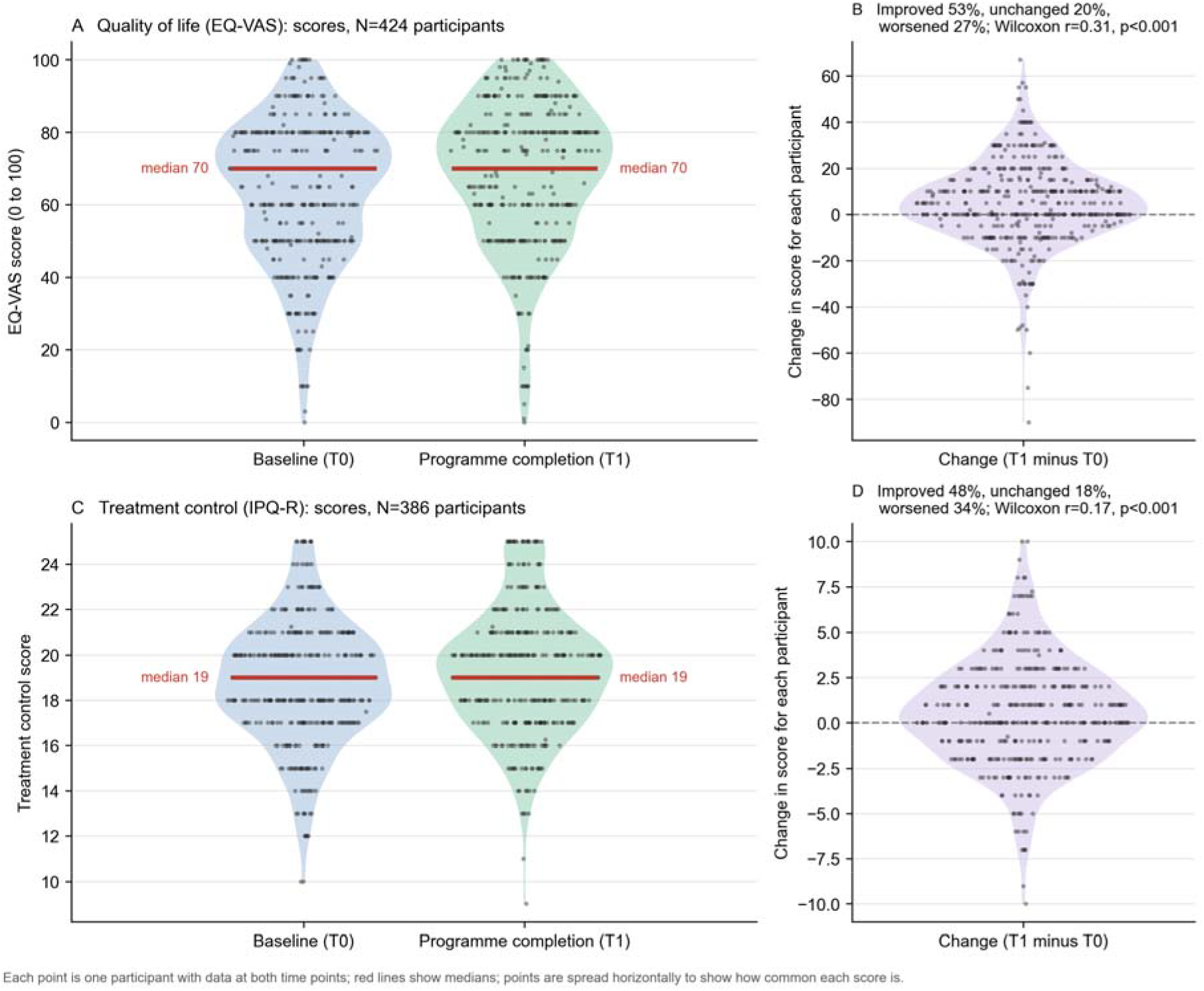
Individual change in quality of life (EQ-VAS) and treatment control (IPQ-R) from baseline to programme completion. *Panels A and C show scores at baseline (T0) and programme completion (T1) for participants with data at both ti points (quality of life, N=424; treatment control, N=386, UK, France and Belgium only). Each point is one participant, red lines show medians, and points are spread horizontally to show how common each score is. Panels B and D show each participant’s change in score (T1 minus T0), with the dashed line marking no change. Although the median score did not change for either outcome, more participants improved than worsened, which the paired Wilcoxon signed-rank test detects (quality of life: 53% improved, 27% worsened, r=0*.*31, p<0*.*001; treatment control: 48% improved, 34% worsened, r=0*.*17, p<0*.*001)*.

Compared with baseline, most patient-reported outcomes remained significantly different at six-month follow-up (Table 4), including empowerment (median 28 to 31; r=0.50), illness coherence (16 to 20; r=0.56), emotional representations (18 to 16; r=0.32), emotional eating (34 to 28; r=0.26) and external eating (27 to 25; r=0.23), all p<0.001. Treatment control, perceived timeline and perceived consequences did not differ significantly from baseline at follow-up.

#### Empowerment and illness perceptions

Empowerment rose significantly from baseline to programme completion (N=414, r=0.63, p<0.001). Between programme completion and six-month follow-up there was a small but statistically significant decline (N=255, r=0.14, p<0.05), with the median falling from 32.0 to 31.0. Empowerment at follow-up nevertheless remained well above the baseline median of 28.0, indicating that most of the gain achieved during the programme was retained.

Positive illness perceptions also improved over the programme. Illness coherence reflecting participants’ understanding of their diabetes rose significantly (N=388, r=0.63, p<0.001), as did treatment control (N=386, r=0.17, p<0.001) and personal control (N=388, r=0.29, p<0.001). Between programme completion and follow-up, only treatment control changed significantly (N=240, r=0.13, p<0.05), and at follow-up it no longer differed significantly from baseline (N=242, r=0.08, p=0.216).

Among the negative illness-perception domains, emotional representations (N=383, r=0.37, p<0.001) and perceptions of diabetes as a cyclical illness (N=382, r=0.13, p<0.05) fell significantly by programme completion. Changes in perceived timeline, acute versus chronic (N=385, r=0.07, p=0.162), and perceived consequences (N=386, r=0.04, p=0.463) did not reach significance, and none of these domains changed further at six-month follow-up.

#### Eating behaviour and health status

By programme completion, restrained eating had increased significantly (N=409, r=0.32, p<0.001), while both emotional eating (N=417, r=0.27, p<0.001) and eating in response to external cues (N=420, r=0.26, p<0.001) had decreased.

Between programme completion and follow-up, restrained eating fell slightly (N=252, r=0.22, p<0.001), though it remained above baseline levels; emotional and external eating did not change further.

The divergence between restrained eating, which increased, and emotional and external eating, which decreased, is worth noting. These constructs are often positively correlated in general population samples, where high dietary restraint frequently co-occurs with emotionally driven eating. The pattern of greater restraint alongside reduced emotional and external eating was observed which is more consistent with the deliberate, planned dietary control that the programme sought to encourage than with the restraint– disinhibition cycle typically seen in unstructured dieting, although this interpretation is tentative and was not tested directly. Increased dietary restraint is not unambiguously beneficial, as it can in some contexts be a precursor to disordered eating, and this warrants monitoring in future evaluations.

Physical health (N=383, r=0.22, p<0.001) and mental health (N=383, r=0.25, p<0.001) both improved significantly from baseline to programme completion, with no further change at six-month follow-up.

### Physical activity (IPAQ)

Physical activity was assessed using the International Physical Activity Questionnaire (IPAQ), with results expressed as median metabolic equivalent of task (MET) minutes per week, in which time spent in each type of activity is weighted by its energy cost relative to rest. These analyses were based on reduced samples, as participants with missing or implausible responses were excluded in line with the instrument’s scoring guidance, and the findings should be read with this in mind.

Total physical activity increased from baseline to programme completion in all four countries. The increase was statistically significant in the UK (1386 to 2040 MET-minutes/week; p=0.005). In France the rise in total activity did not reach significance (1671 to 2226; p=0.080), although walking activity increased significantly (396 to 693; p=0.007). In Belgium (2030 to 3565; p=0.465) and the Netherlands (3036 to 3068; p=0.790) the upward trend was not statistically significant.

### Health-related quality of life (EQ-VAS)

Health-related quality of life was measured using the visual analogue scale (EQ-VAS) from the EQ-5D-3L, on which participants rate their health from 0 (worst imaginable) to 100 (best imaginable). The EQ-5D-3L utility index and quality-adjusted life years are not reported here, as the non-intervention comparison group was not recruited owing to the COVID-19 pandemic. Self-rated quality of life showed a statistically significant improvement from baseline to programme completion (N=424, r=0.31, p<0.001), with no further change by six-month follow-up (N=148, r=0.07, p=0.36). This result requires careful interpretation: the median EQ-VAS score was 70.00 at all three time points, so the significant test statistic reflects a consistent direction of within-person change rather than a shift in the central tendency of the sample. The magnitude of any underlying change in perceived quality of life is therefore likely to be modest, and the finding should not be read as evidence of a substantial improvement.

### Comparison of participants retained at follow-up with those lost

To assess whether the follow-up sample remained representative, participants retained at six-month follow-up (n=293) were compared with those lost (n=312) on baseline characteristics. Continuous variables were compared using Mann–Whitney U tests and categorical variables using chi-square tests, with Benjamini–Hochberg correction applied across all sixteen comparisons to control the false discovery rate (Table 5).

**Table 5.** Baseline characteristics of participants retained at six-month follow-up versus those lost.

| **Baseline characteristic** | **Retained, median [IQR]** | **Lost, median [IQR]** | **p (raw)** | **p (adj.)** |
|---|---|---|---|---|
| Age | 64.0 [58–69] | 62.3 [55–69] | 0.040 | 0.129 |
| Time since diagnosis (yrs) | 9.0 [4–15] | 10.0 [3–15] | 0.600 | 0.686 |
| Weight (kg) | 95.0 [82–109] | 94.4 [80–112] | 0.840 | 0.840 |
| BMI (kg/m²) | 33.9 [30–38] | 32.6 [29–38] | 0.150 | 0.343 |
| Waist (cm) | 112.0 [103–122] | 111.0 [100–123] | 0.480 | 0.663 |
| HbA1c (mmol/mol) | 56.1 [48–68] | 59.9 [49–70] | 0.061 | 0.163 |
| Empowerment (DES-SF) | 28.0 [24–31] | 28.0 [25–31] | 0.330 | 0.659 |
| Illness coherence | 16.0 [14–20] | 16.0 [12–19] | 0.020 | 0.109 |
| Restrained eating | 28.0 [24–33] | 29.0 [24–33] | 0.474 | 0.663 |
| Emotional eating | 33.0 [22–43] | 34.0 [23–44] | 0.501 | 0.663 |
| Physical health (SF-12) | 44.8 [35–51] | 39.9 [31–50] | 0.005 | 0.044* |
| Mental health (SF-12) | 45.6 [35–53] | 44.4 [35–54] | 0.538 | 0.663 |
| Quality of life (EQ-VAS) | 70.0 [50–80] | 65.0 [50–80] | 0.039 | 0.129 |
| Sex | — | — | 0.425 | 0.663 |
| Ethnicity | — | — | 0.781 | 0.833 |
| Country | — | — | <0.001 | <0.001* |

*Continuous variables: Mann–Whitney U; categorical: chi-square. p (adj.) = Benjamini–Hochberg adjusted. \*significant after correction.*

After correction, the two groups differed on only two characteristics. Retention differed by country (adjusted p<0.001), reflecting the COVID-19 disruption that particularly affected the Belgian cohort (retention by country: UK 44%, France 63%, Netherlands 46%, Belgium 17%). Those retained also had modestly better baseline physical health (SF-12; adjusted p=0.044), suggesting that participants in poorer physical health were somewhat more likely to be lost to follow-up. Baseline HbA1c did not differ significantly after correction (retained 56.1 vs lost 59.9 mmol/mol; raw p=0.061, adjusted p=0.163), though the direction of poorer glycaemic control among those lost is consistent with the physical-health finding. All other baseline characteristics, including age, sex, ethnicity, anthropometry, and the remaining psychological measures, were comparable between groups, supporting the broad representativeness of the follow-up sample.

### Referral routes

Referral route was recorded for 570 of 605 participants (94%). Routes differed substantially between countries and were not defined consistently across sites, limiting direct comparison (Table 6). In the UK, most participants self-referred (45%) or entered via routes recorded as “other” (37%), with 17% referred by a GP. In France, “other” routes (58%) and hospital referral (30%) predominated. Recruitment in the Netherlands was almost entirely hospital-based (98%), and in Belgium was chiefly by self-referral (59%) or “other” routes (41%). The high proportion coded “other” in several countries indicates that the referral categories, which reflected UK service structures, did not map cleanly onto other national pathways. In Belgium, for example, participants were recruited largely through mutual health insurers (via their online and offline communication channels) and community health centres, often by a diabetes nurse rather than a GP.

**Table 6.** Referral route by country.

| **Country** | **GP surgery** | **Hospital** | **Self-referral** | **Other** | **Recorded, n** |
|---|---|---|---|---|---|
| UK | 47 (17%) | 2 (1%) | 121 (45%) | 100 (37%) | 270 |
| France | 13 (7%) | 60 (30%) | 11 (6%) | 117 (58%) | 201 |
| Netherlands | 0 | 54 (98%) | 1 (2%) | 0 | 55 |
| Belgium | 0 | 0 | 26 (59%) | 18 (41%) | 44 |

*Percentages are within-country, of participants with a recorded referral route.*

## Discussion

Participation in DWELL was linked to short-term improvements across metabolic, psychological and self-management outcomes. Weight, BMI, waist circumference and HbA1c all improved after completion of the twelve-week programme, alongside gains in empowerment, illness understanding, perceived control, eating behaviour, and physical and mental health; improvements in self-care and medication adherence were also observed, though these should be interpreted with some caution, as the SDSCA responses required harmonisation across time points before analysis. Taken together, these findings make the case for evaluating DSMES programmes on both clinical and patient-reported outcomes, rather than on glycaemic control alone.

The metabolic findings sit comfortably alongside existing evidence that structured diabetes education and support can improve glycaemic control and other clinical outcomes.^5–7^ For example, a network meta-analysis of 108 randomised trials found that combined self-management education and support reduced HbA1c more than usual care or education alone,^6^ and a systematic review of 44 studies reported HbA1c reductions of 0.64% in randomised trials and 1.27% in quasi-experimental studies.^7^ Yet HbA1c rose again between programme completion and follow-up, although it remained below its baseline level (a median of 1.7 mmol/mol lower at follow-up), echoing a familiar pattern in the literature: gains tend to be largest during or shortly after active programme delivery, and to fade once structured contact ends.^8,9^ A meta-review of 41 systematic reviews, drawing on 459 randomised trials across 33 countries, found HbA1c improvements of 0.2 to 0.6% (2.2 to 6.5 mmol/mol) at six months that attenuated progressively by 12 and 24 months, with multicomponent programmes offering at least 10 hours of contact faring best,^8^ and dose-response evidence suggests a similar drift back towards baseline after around 12 months.^9^ This does not necessarily mean that education is ineffective in the longer term; it more likely reflects a need for ongoing reinforcement and opportunities to revisit self-management skills if early gains are to be sustained. In this study that pattern may have been compounded by the COVID-19 pandemic, which interrupted delivery and follow-up across all four countries and restricted access to routine care and community-based physical activity during the period covered by the 6-month assessments. Several psychological and behavioural outcomes, by contrast, held up better over the same period suggesting that the confidence, understanding and engagement participants gained may outlast the metabolic effects.

The improvements in empowerment, illness coherence and perceived personal and treatment control are worth dwelling on, because effective self-management depends not just on what people know but on their confidence to act on that knowledge day to day. This fits with earlier evidence that person-centred approaches built around goal setting, problem-solving, social support and self-efficacy can shift these outcomes.^23,24^ A systematic review of illness-perception interventions found that illness coherence, personal control and treatment control were the perceptions most open to change, although improving them did not consistently improve HbA1c,^23^ and a large patient-centred self-management trial reported substantial gains in self-efficacy and self-care behaviours alongside a smaller improvement in HbA1c, suggesting that behavioural and psychological change can occur independently of, or ahead of, metabolic change.^24^ The present pattern, in which psychological gains were largely retained while the HbA1c improvement partly faded, is consistent with this. Diabetes education and empowerment are thought to strengthen people’s sense of control over their condition and their self-care,^25^ and perceived locus of control and self-control are recognised drivers of self-management behaviour in long-term conditions, although they are rarely assessed directly in programme evaluations;^26,27^ a stronger internal locus of control and greater self-control have been associated with better physical health and psychological wellbeing.^28^ The reductions in emotional and external eating, together with the gains in physical and mental health, add further weight to the case for building behavioural and psychosocial components into diabetes education. This matters because diabetes distress, depression and anxiety are linked to poorer self-care and glycaemic outcomes,^29^ and tailored psychological interventions can reduce diabetes distress even where HbA1c does not change.^30^ That said, this study did not test directly whether changes in empowerment or illness perceptions drove the metabolic improvements observed.

Two features of the analysis temper these conclusions. No correction was applied to the longitudinal outcome comparisons, so with the number of outcomes and time points examined some significant results may be chance findings; and several of the significant changes, particularly between programme completion and follow-up, were small in magnitude with correspondingly small effect sizes. Statistical significance in a sample of this size does not by itself establish clinical importance. The findings are best treated as hypothesis-generating, indicating where a controlled evaluation might most usefully focus rather than establishing effect sizes that could be relied upon for planning.

There are also implications here for how such programmes are designed. The fading HbA1c benefit suggests that 12 weeks may be enough to set change in motion but not always enough to sustain it without further reinforcement. Future work should test whether booster sessions, peer support, personalised follow-up or structured opportunities to re-engage can help outcomes last longer. Controlled studies with longer follow-up are also needed to establish the programme’s effectiveness more robustly, determine which components of the programme matter most, and examine whether outcomes differ by country, level of participation, ethnicity, socioeconomic status or health literacy. Barriers to self-management are rarely singular: health literacy, communication with healthcare professionals, socioeconomic circumstances, comorbidities, cultural context and the availability of family, peer and professional support can all play a part,^31–33^ and many people do not engage effectively with self-management early after diagnosis, particularly those facing compounding factors such as serious mental illness or multiple deprivation.^8^ People also differ in their readiness to change long-established habits: some fold new behaviours into daily life relatively easily, while others struggle to reconcile what they want to do with what they feel they should do, or disengage altogether.^34^ Community-based, culturally sensitive programmes that combine incremental learning, hands-on practice, peer support and personalised feedback appear well placed to address these barriers,^33,35^ which is what DWELL’s flexible, person-centred design sought to do. Delivery across four countries also highlights the need to adapt such programmes to local services, cultures and populations: the Diabetes Literacy project found that only around 29% of respondents across European Union member states reported DSME programmes adapted for people with limited literacy, education or language proficiency, compared with 63% in comparator countries outside the EU.^10^ Data from the Dutch DWELL cohort have also been used to examine functional outcomes: in a cross-sectional analysis combining DWELL participants with a second cohort, peripheral neuropathy was associated with poorer daily-life gait quality, including slower walking speed,^36^ suggesting that future evaluations could usefully assess functional outcomes alongside clinical and patient-reported measures. There is also scope to explore the cultural and intersectional dimensions of self-management support more directly including how programmes like DWELL might be adapted for people managing T2DM alongside significant comorbidities such as serious mental illness.

### Limitations

These findings should be read considering the study’s uncontrolled, pre–post design, which means the changes observed cannot be attributed to the DWELL programme alone. Without a comparison group, secular trends, regression to the mean and the effects of repeated assessment cannot be ruled out.

Programme delivery in Belgium was interrupted at all three sites and did not resume after the pandemic; although new delivery sites were sought and a further recruitment campaign was undertaken, participant numbers were insufficient to restart the programme. This contributed substantially to the pronounced attrition at that site.

This disruption has several implications. People with T2DM were advised to shield, group-based sessions were suspended, face-to-face assessment was interrupted, and access to routine diabetes care and to the community-based physical activity options that formed part of the programme was reduced. These circumstances are likely to have contributed to the loss to follow-up seen at T2 and may have compounded the attenuation of glycaemic improvement observed between programme completion and follow-up, alongside the expected effect of structured contact ending. The pandemic is also likely to have curtailed recruitment, as group delivery was suspended and many potential participants, who were often older and advised to shield, may have been reluctant to join group-based programmes. Beyond these practical effects, the pandemic may have changed how participants viewed their diabetes and their self-care. People with diabetes were identified early as being at higher risk of severe COVID-19, and studies from this period reported widespread diabetes-specific worries, including fear of being badly affected by the virus and of being unable to manage diabetes if infected,^37^ as well as increased stress and anxiety, weight gain and reduced exercise during lockdown.^38^ Such experiences could have shifted participants’ illness perceptions, emotional responses to diabetes and self-care behaviours at follow-up in either direction, strengthening motivation for self-management in some participants while undermining it in others, and the present data cannot separate these influences from the effects of the programme. Because cohorts were affected at different stages of their programme, and to differing degrees between countries, the pandemic represents an unmeasured source of heterogeneity that this analysis cannot isolate. Retention was, however, markedly lower in Belgium than elsewhere, consistent with this disruption.

The reduction in participant numbers by follow-up is a further consideration. Participants retained at follow-up differed modestly from those lost in baseline physical health, those retained being somewhat healthier (see Comparison of participants retained at follow-up with those lost). Follow-up outcomes may therefore be modestly favourable through selective retention, and baseline HbA1c showed a non-significant difference in the same direction. The two groups were otherwise comparable, and no other baseline characteristic differed after correction. More broadly, a self-selecting, self-management programme is likely to attract more motivated participants, and reaching and recruiting less motivated people with T2DM was a persistent challenge; further research is needed to establish whether the programme benefits people with T2DM generally or is more effective among those who are already highly motivated, which may also have influenced the present results. A related limitation is the absence of correction for multiple comparisons in the longitudinal outcome analyses: given the number of outcomes and repeated time points examined, some significant findings may represent type I errors, and the results are best regarded as hypothesis-generating. The primary analyses were unadjusted; however, the covariate-adjusted models (Supplementary Table S1) indicate that the programme effects on the key metabolic and psychological outcomes persisted after adjustment for demographic characteristics and country of delivery.

Finally, the programme was delivered across four countries and healthcare settings, and local differences in implementation and service context may have shaped outcomes in ways this analysis cannot reveal. These factors limit how far the results can be generalised or interpreted causally, but they do not lessen the value of this study as a large, real-world, multinational evaluation of a complex diabetes self-management programme delivered under demanding conditions.

## Conclusions

Participation in the 12-week DWELL programme was associated with improvements across metabolic, psychological and self-management outcomes in adults with T2DM across four European countries.

Many of these gains held at six-month follow-up, most clearly for weight, waist circumference, empowerment, illness coherence, and self-care and adherence behaviours, although the early improvement in HbA1c had partly faded by then, while remaining below baseline. Together, the findings support the value of flexible, person-centred DSMES models that pair education with behavioural and psychosocial support. Because the study used an uncontrolled design and experienced loss to follow-up, however, the results cannot establish causation. Controlled evaluations are now needed to test how ongoing support, programme fidelity and local context shape how durable these clinical and patient-reported benefits prove to be.

## Data Availability

All data produced in the present study are available upon reasonable request to the authors

## Acknowledgements

We are grateful to all DWELL participants in the four countries for their time and contribution, and to the people living with diabetes, family carers, healthcare professionals and academics who co-designed the programme. We thank the partner organisations responsible for programme delivery and data collection, namely Medway Community Healthcare and Blackthorn Trust in the UK, Artevelde University of Applied Sciences in Belgium, Centre Hospitalier de Douai in France and Kinetic Analysis in the Netherlands, and in particular the following individuals: in the UK, Julie Webster, Anne Eltringham-Cox and Jane Redding (Medway Community Healthcare), Nathalie Belmas and Sue Shaw (Blackthorn Trust), and Stephen Cochrane (Kent County Council); in Belgium, Annelien Callens and Veerle Luyens (Artevelde University of Applied Sciences); in France, Marie Duezcalzada, Jerome Cazier and Dr Véronique Averous (Centre Hospitalier de Douai); and in the Netherlands, Linda van Wijk and Melvin Franken (Kinetic Analysis). The authors acknowledge with thanks the contributions of Rachael Morris, Thomas Thompson, Julia Moore and David Vernon, who contributed to an earlier draft of this work. Although the manuscript has subsequently been substantially revised and developed, their original contributions helped shape the work presented here. We also thank Dr Marlize De Vivo and Prof Kate Springett (Canterbury Christ Church University) and Dr Katrina Taylor (University of Kent) for their contributions at earlier stages of the evaluation. Finally, we thank Anna Best and Alice Chapman-Hatchett, members of the original project management team, for clarifying the project, addressing revision queries and facilitating access to relevant data and documentation, and Dr Ben Field (Associate Professor of Diabetes and Endocrinology, University of Surrey) for his helpful comments on the manuscript.

