## Supporting Information for "Participant Outcomes of the Diabetes and WELLbeing 12-Week Psychoeducational Self-Management Programme for Type 2 Diabetes: A Four-Country European Evaluation"

### Supplementary Material

#### **Effect of adjustment for demographic and country characteristics**

To test whether the programme effects were robust to participant characteristics, covariate-adjusted models were fitted for key metabolic and psychological outcomes. Each outcome was modelled from baseline to programme completion using linear regression with cluster-robust standard errors by participant, which account for the two repeated measures per person. Models were adjusted for age, sex, time since diagnosis, country, and baseline physical health (except where physical health was the outcome). A site random intercept was considered but was singular: with only five site groupings, nearly nested within country, the country fixed effect captured the between-site variation, so the random intercept was not retained (Supplementary Table S1).

**Supplementary Table S1 Covariate-adjusted programme effects (baseline to programme completion) for key metabolic and psychological outcomes.**

| **Outcome** | **Adjusted effect** | **95% CI** | **p** | **N (obs / participants)** |
| --- | --- | --- | --- | --- |
| HbA1c (mmol/mol) | −3.25 | −4.58 to −1.92 | <0.001 | 681 / 394 |
| Weight (kg) | −1.84 | −3.06 to −0.63 | 0.003 | 731 / 414 |
| Waist circumference (cm) | −2.15 | −3.16 to −1.15 | <0.001 | 705 / 404 |
| Empowerment (DES-SF) | +3.79 | +3.20 to +4.37 | <0.001 | 731 / 415 |
| Physical health (SF-12) | +2.47 | +1.43 to +3.51 | <0.001 | 743 / 430 |

*Linear regression with cluster-robust standard errors by participant, adjusted for age, sex, time since diagnosis, country and baseline physical health. Participant counts exceed the paired within-person Ns reported for the Wilcoxon analyses because the repeated-measures models retain participants contributing data at only one time point.*

All five outcomes retained significant improvements in the expected direction after adjustment. HbA1c and weight fell, and waist circumference, empowerment and physical health improved. Adjustment for demographic and country characteristics therefore did not alter the programme’s apparent effects. The three anthropometric outcomes (weight, BMI and waist circumference) were closely correlated and represent one metabolic dimension rather than independent confirmations.
